# Outcome of hospitalisation for COVID-19 in patients with Interstitial Lung Disease: An international multicentre study.

**DOI:** 10.1101/2020.07.15.20152967

**Authors:** Thomas M Drake, Annemarie B Docherty, Ewen M Harrison, Jennifer K Quint, Huzaifa Adamali, Sarah Agnew, Suresh Babu, Christopher M Barber, Shaney Barratt, Elisabeth Bendstrup, Stephen Bianchi, Diego Castillo Villegas, Nazia Chaudhuri, Felix Chua, Robina Coker, William Chang, Anjali Crawshaw, Louise E. Crowley, Davinder Dosanjh, Christine A Fiddler, Ian A. Forrest, Peter George, Michael A Gibbons, Katherine Groom, Sarah Haney, Simon P Hart, Emily Heiden, Michael Henry, Ling-Pei Ho, Rachel K Hoyles, John Hutchinson, Killian Hurley, Mark Jones, Steve Jones, Maria Kokosi, Michael Kreuter, Laura MacKay, Siva Mahendran, George Margaritopoulos, Maria Molina-Molina, Philip L Molyneaux, Aiden O’Brien, Katherine O’Reilly, Alice Packham, Helen Parfrey, Venerino Poletti, Joanna Porter, Elisabetta Renzoni, Pilar Rivera-Ortega, Anne-Marie Russell, Gauri Saini, Lisa G Spencer, Giulia M. Stella, Helen Stone, Sharon Sturney, David Thickett, Muhunthan Thillai, Tim Wallis, Katie Ward, Athol U Wells, Alex West, Melissa Wickremasinghe, Felix Woodhead, Glenn Hearson, Lucy Howard, J Kenneth Baillie, Peter J.M. Openshaw, Malcolm G Semple, Iain Stewart, ISARIC4C Investigators, R Gisli Jenkins

## Abstract

**Rationale:** The impact of COVID-19 on patients with Interstitial Lung Disease (ILD) has not been established.

**Objectives:** To assess outcomes following COVID-19 in patients with ILD versus those without in a contemporaneous age, sex and comorbidity matched population.

**Methods:** An international multicentre audit of patients with a prior diagnosis of ILD admitted to hospital with COVID-19 between 1 March and 1 May 2020 was undertaken and compared with patients, without ILD obtained from the ISARIC 4C cohort, admitted with COVID-19 over the same period. The primary outcome was survival. Secondary analysis distinguished IPF from non-IPF ILD and used lung function to determine the greatest risks of death.

**Measurements and Main Results:** Data from 349 patients with ILD across Europe were included, of whom 161 were admitted to hospital with laboratory or clinical evidence of COVID-19 and eligible for propensity-score matching. Overall mortality was 49% (79/161) in patients with ILD with COVID-19. After matching ILD patients with COVID-19 had higher mortality (HR 1.60, Confidence Intervals 1.17-2.18 p=0.003) compared with age, sex and comorbidity matched controls without ILD. Patients with a Forced Vital Capacity (FVC) of <80% had an increased risk of death versus patients with FVC ≥80% (HR 1.72, 1.05-2.83). Furthermore, obese patients with ILD had an elevated risk of death (HR 1.98, 1.13−3.46).

**Conclusions:** Patients with ILD are at increased risk of death from COVID-19, particularly those with poor lung function and obesity. Stringent precautions should be taken to avoid COVID-19 in patients with ILD.

## Introduction

Interstitial Lung Diseases (ILDs) represent a group of fibroinflammatory diseases affecting the alveolar interstitium of the lung. ILDs are characterised by alveolar damage and interstitial thickening and, if left untreated, lead to remorseless progression of breathlessness, cough and ultimately death from respiratory failure. The prevalence of ILDs in Europe is just under one per 1,000 people, with an annual incidence of approximately 20 per 100,000 people (1). The commonest ILD is sarcoidosis, but one of the most severe ILD is Idiopathic Pulmonary Fibrosis (IPF), a high incidence of which is found in the UK (12 per 100,000) (2, 3). IPF tends to affect older people with a mean age at diagnosis of 72 years. Men are more often affected than women. Furthermore, IPF is associated with diabetes mellitus type 2 (DMT2), hypertension (HT) and ischaemic heart disease (IHD) (4, 5). People suffering from other ILDs (non-IPF ILD) tend to be younger, with a higher proportion of female sufferers, and often receive immunosuppressive therapy.

All ILDs, most notably IPF, are characterised by acute exacerbations which have a particularly high mortality rate ranging from 35-70% (6). The precise cause of acute exacerbations is unknown, but they have been associated with thoracic surgical procedures and viral infections (6). Furthermore, an acute exacerbation is associated with the development of Acute Respiratory Distress Syndrome (ARDS) which carries a high mortality and morbidity. However, there is no consensus on treatment of ARDS in this group of patients, given that mechanical ventilation is the cornerstone of supportive therapy in non-ILD patients (7).

Infection with SARS-CoV-2 may lead to COVID-19, characterised by a severe viral pneumonia and ARDS in approximately 20% of patients admitted to hospital (8). Patients most at risk of severe COVID-19 include elderly males with comorbidities including DMT2, HT and IHD (9,10) which are shared by patients with ILD, most notably IPF (5). Despite the potential increased risk from SARS-CoV-2 infection, patients with ILD have not been uniformly advised to isolate themselves from the rest of the population to avoid infection at the onset of the COVID-19 pandemic.

To understand whether individuals with ILD are more susceptible to severe COVID-19 and should therefore be advised to self-isolate during the pandemic, we undertook an international multicentre analysis of patients admitted to hospital between the 1 March and 1 May 2020 with COVID-19. We compared outcomes of patients with ILD with age, sex and comorbidity matched controls admitted with COVID-19 but without ILD, from a prospective UK cohort, the International Severe Acute Respiratory and Emerging Infection Consortium Coronavirus Clinical Characterisation Protocol (ISARIC4C CCP-UK).

## Methods

### Patients

ILD centres throughout Europe were asked to identify all patients with a pre-existing diagnosis of ILD admitted to hospital between 1st March and 1st May 2020 during the first peak of the COVID-19 pandemic. De-identified, unlinked data from individuals were included in an audit to assess the consequence of national shielding policies. Each contributing site was asked to identify individuals admitted to their local hospital. Local hospitals were asked to include whether or not a patient was diagnosed with COVID-19 based on a SARS-CoV-2 positive polymerase chain reaction (PCR) swab and or clinicoradiological diagnosis for patient flow (Figure 1). Audit data was appended to the ISARIC4C CCP-UK database, with separate categorisation of individuals who reported an existing chronic pulmonary disease. Significant comorbidities of diabetes (type 1 or type 2), hypertension, chronic heart disease and malignancy, as well as immunomodulatory therapies, were recorded in both ILD audit data and the ISARIC database. Outcomes recorded in both datasets for each patient included ventilated status, use of continuous positive airway pressure, use of high flow oxygen and whether discharged, remained hospitalised, or died. Length of stay was censored at 1st May 2020.

**Figure 1.**
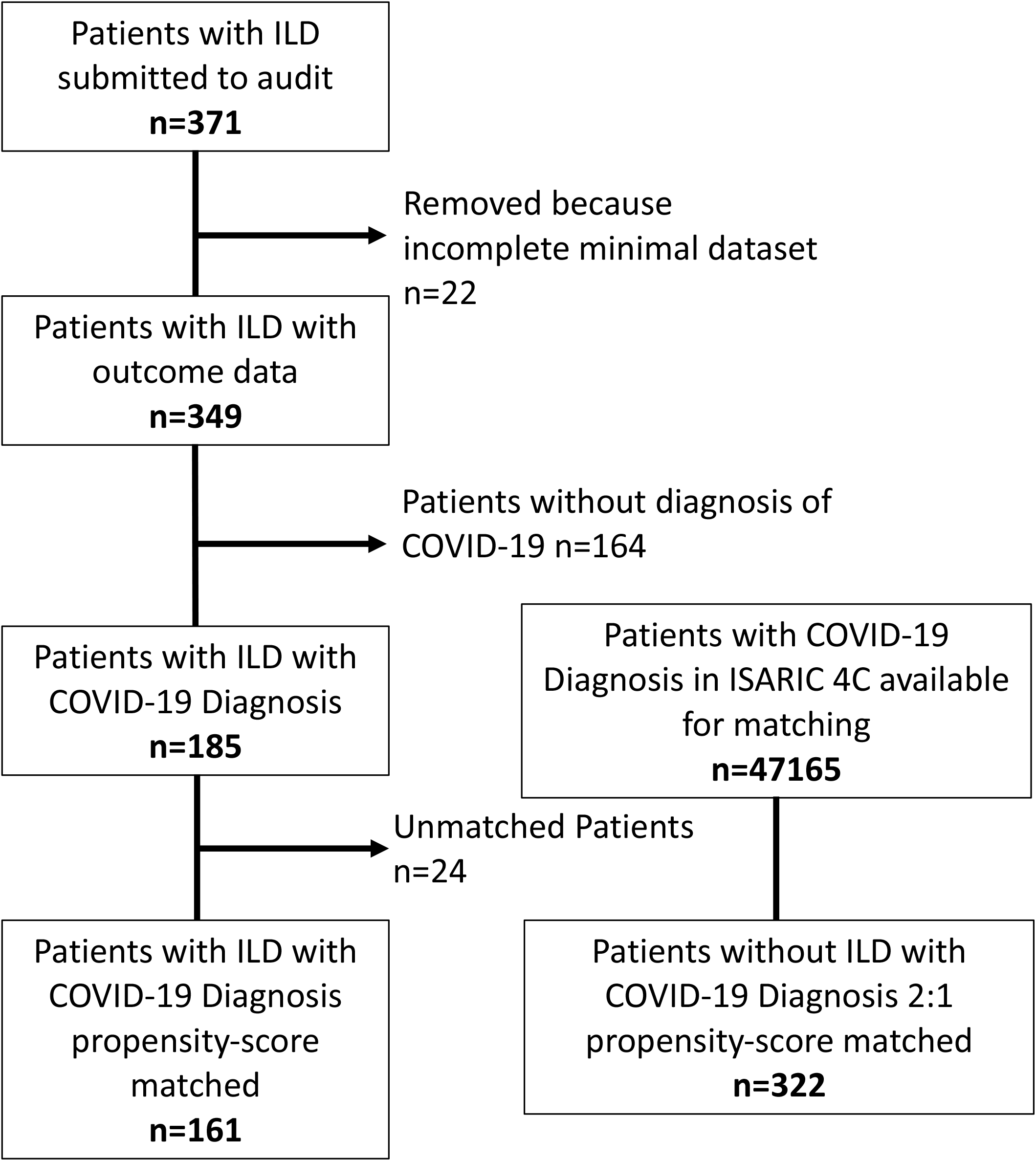
CONSORT Diagram showing patients recruited and flow to matching.

### Ethics and Consent

All data were entered by the Local Clinical Care Team in anonymised fashion without linkage to any patient identifiers in line with national and local audit guidance. In the UK Health Research Authority guidance was followed (https://www.hra.nhs.uk/covid-19-research/guidance-using-patient-data/), and ethical approval was not required. ISARIC4C CCP-UK received ethical approval from the South Central – Oxford C Research Ethics Committee in England (Ref: 13/SC/0149), and by the Scotland A Research Ethics Committee (Ref: 20/SS/0028). Data from Spain was collected under the approval for observational studies (Ref UIC-IBU-2020-03 and PR217/20). Data from Germany were collected under approval from ethics committee of the Medical faculty of the University of Heidelberg (ref S-186/2020).

### Statistical Analysis

Summary statistics are presented as frequencies and percentages for categorical data and mean (standard deviation (sd)) for normally distributed continuous data. Where continuous data were not normally distributed, the median (interquartile range [IQR]) was used.

Differences in categorical data were compared using the Chi-square test, or Fisher’s exact test when expected counts were below 5 in any group. For continuous normally distributed 2-group data, we compared differences using Welch’s T-test or Mann Whitney-U if data were not normally distributed. For multiple group comparisons of continuous data, we used Kruskall-Wallis tests. Data from the ILD dataset were matched in a 1:2 ratio with patients who did not have ILD from the ISARIC4C CCP-UK dataset using a nearest neighbour propensity-score matching algorithm. Patients in ISARIC4C CCP-UK with ILD were excluded to avoid double counting. We matched on confounders known to affect outcomes from COVID-19 disease including age, sex, diabetes, chronic cardiac disease, cancer, hypertension, and renal disease. Unmatched and matched populations were compared using standardized mean differences, plots and summary statistics. For survival analyses, time was taken as admission to death (in-hospital survival) using length of stay where date was not available. Discharge from hospital was considered an absorbing state (once discharged, patients were considered no longer at risk of death). Discharged patients were not censored and included in the risk set until the end of follow-up, thus discharge did not compete with death. We generated subclasses based on the propensity-score distance and included these as terms within Cox proportional hazards models, where estimates are presented as hazards ratios, alongside the corresponding 95% confidence interval (11). Data were analysed using R version 3.6.3 with tidyverse, matchit and finalfit packages.

## Results

### Baseline Demographics

Between March 1^st^ and May 1^st^ 2020, 349 patients with ILD were admitted to 37 hospitals throughout the UK and Europe. A total of 185 ILD patients had a diagnosis of COVID-19 and 161 were suitable for propensity-score matching (Figure 1). Of these, 110 (68%) were male and the mean age was 73.2 (±11.5) years. The most common ILD was IPF, with 68 (42%) patients admitted to hospital (Table 1). During the same time period, 164 patients with ILD were admitted with an alternative non-COVID diagnosis, of whom 69 (42.1%) had IPF. Overall the Case Fatality Rate (CFR) for patients with ILD and COVID-19 was 49% (79/161) compared with 17% (28/164) for patients with ILD admitted for other reasons.

**Table 1.**
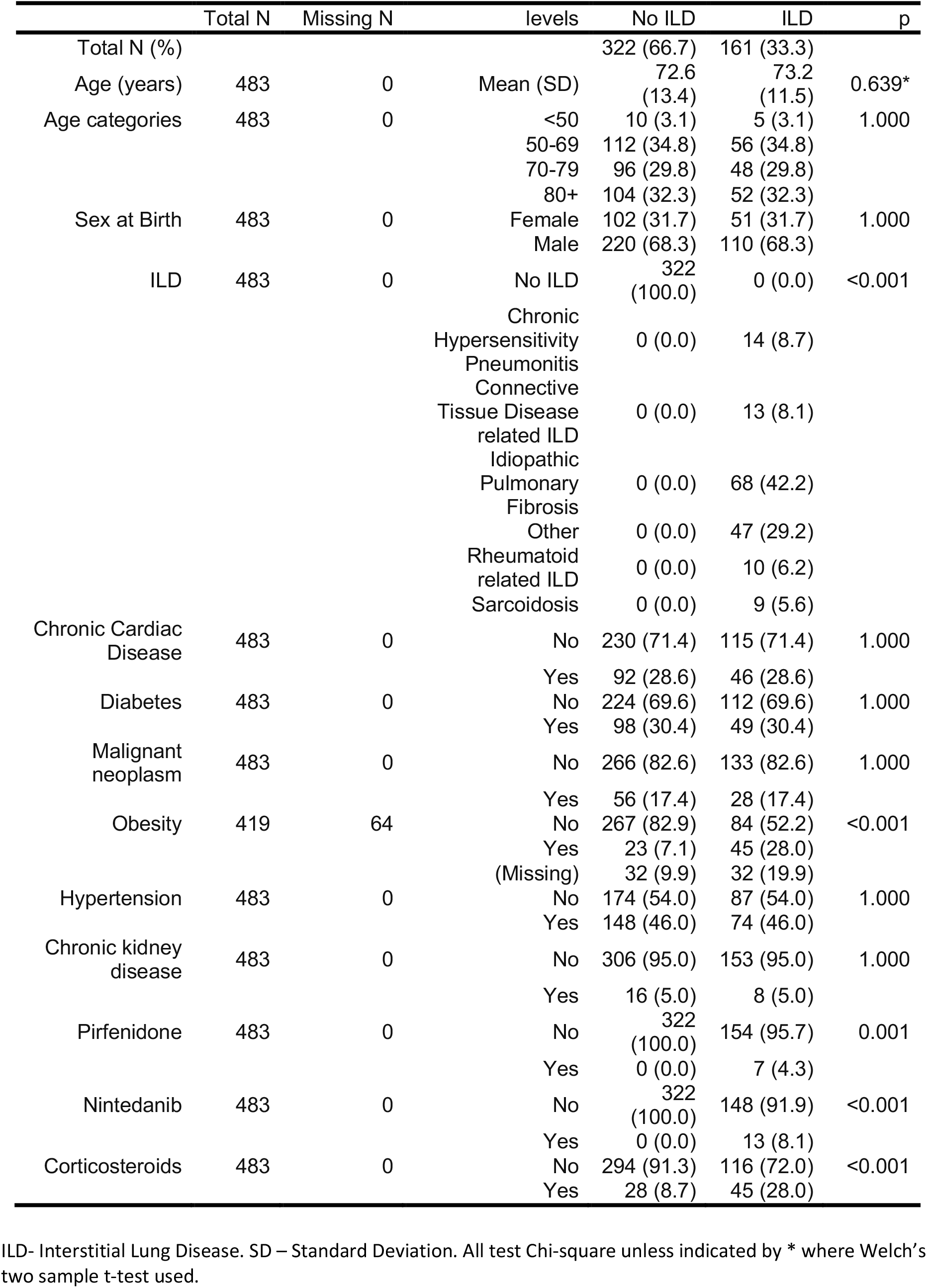
Patient Characteristics by ILD

### Effect of ILD on outcome from COVID-19

After propensity-score matching, 161 patients with ILD were compared with 322 patients admitted to hospital between the 1^st^ March and 1^st^ May 2020 but without ILD, or other chronic lung disease (Table 1). There was significantly higher mortality in patients with ILD compared with non ILD patients (49% (79/161) vs 35% (114/322) p=0.013) and was associated with an increased risk of death in adjusted analysis (HR 1.60 CI 1.17-2.18 p=0.003). There was significantly higher mortality in patients with ILD compared with non ILD patients (figure 2), which was greatest in men and increased with age, which persisted for men after adjusting for age and comorbidity (adjusted HR 1.98, 95% CI 1.14 to 3.43, p = 0.015). The risk of mortality was greatest in patients with IPF (HR 1.74, 95%CI 1.16 to 2.60 p=0.007), but a higher mortality was also seen in non-IPF ILD (HR 1.50, 95%CI 1.02 to 2.21 p=0.040; Figure 3a) when compared with matched patients without ILD. Of patients with non-IPF ILD, those with chronic hypersensitivity pneumonitis and rheumatoid ILD had the highest mortality (50% (7/14) and 40% (4/10) respectively), whilst those with sarcoidosis and connective tissue disease (excluding rheumatoid) related ILD had the lowest observed mortality (33% (3/9) and 23% (3/13) respectively). Overall, median length of stay in those who were still alive was not substantially different between patients without ILD (9 days [IQR 11])), compared with non-IPF ILD (10 days [IQR 8])) and IPF (10 days [IQR 8]) (p = 0.725) (Table 2).

**Table 2:** Outcomes by ILD

|  | Total N | Missing N | levels | No ILD | ILD | p |
| --- | --- | --- | --- | --- | --- | --- |
| Died | 486 | 0 | Alive | 208 (64.6) | 82 (50.9) | 0.013 |
|  |  |  | Died | 114 (35.4) | 79 (49.1) |  |
| Respiratory Support | 486 | 0 | Unenhanced respiratory support* | 254 (78.9) | 136 (84.5) | 0.288 |
|  |  |  | Enhanced Respiratory Support | 39 (12.1) | 20 (12.4) |  |
|  |  |  | Ventilation | 29 (9.0) | 6 (3.7) |  |
| Length of stay for those alive | 290 | 196 | Median (IQR) | 9.0 (11.0) | 10.0 (8.0) | 0.725** |

IQR – Interquartile range. All test Chi-square unless indicated by \*Standard flow-rate Oxygen supplementation not considered ‘enhanced’ respiratory support, CPAP and High Flow O<sub>2</sub> considered enhanced respiratory support. \*\*where Kruskal Wallis used.

**Figure 2.**
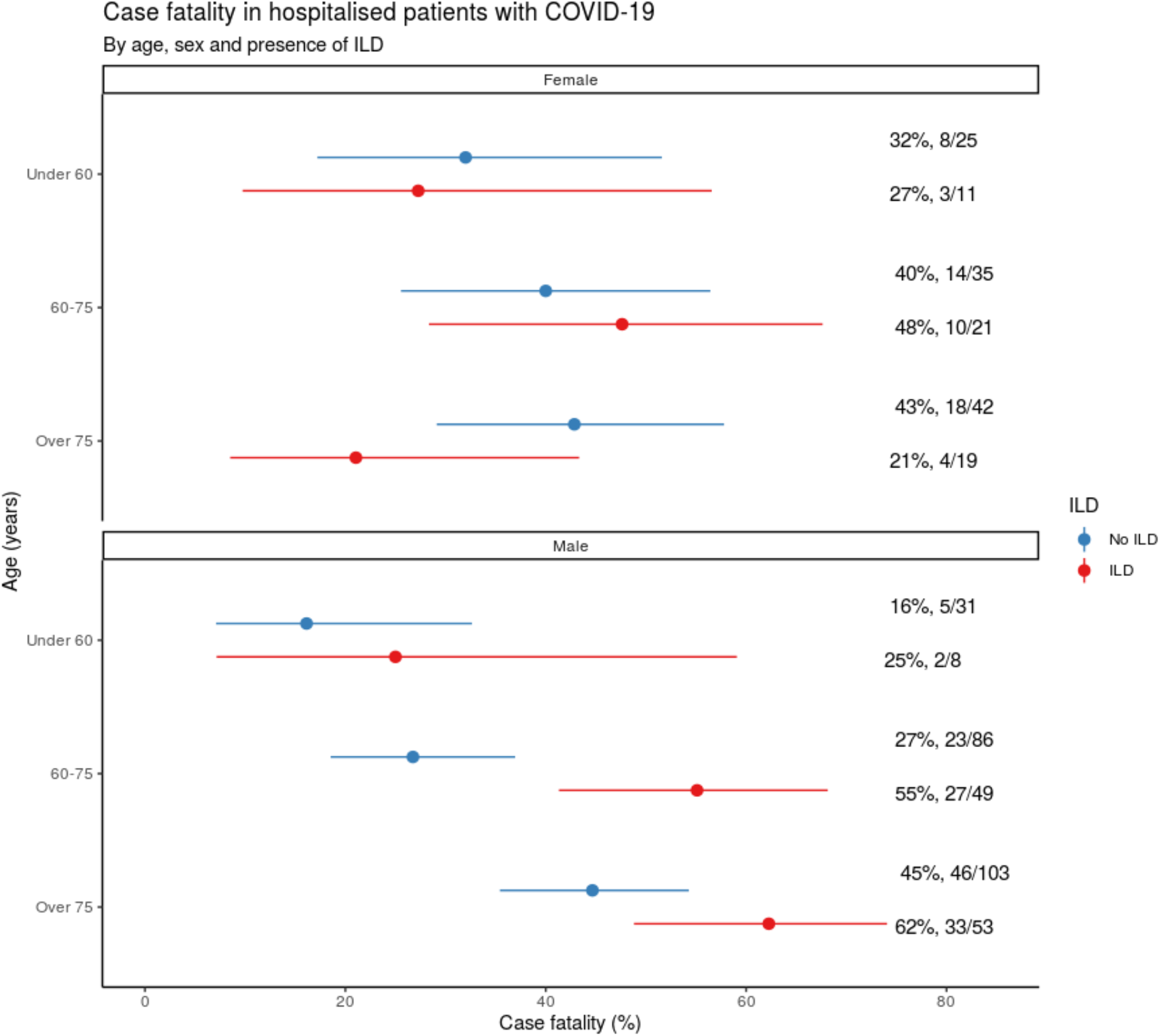
A) Case Fatality Rates with 95% confidence intervals for female ILD patients hospitalised with COVID-19 stratified by age compared with those without ILD. B) Case Fatality Rates with 95% confidence intervals for male ILD patients hospitalised with COVID-19 stratified by age compared with those without ILD.

**Figure 3.**
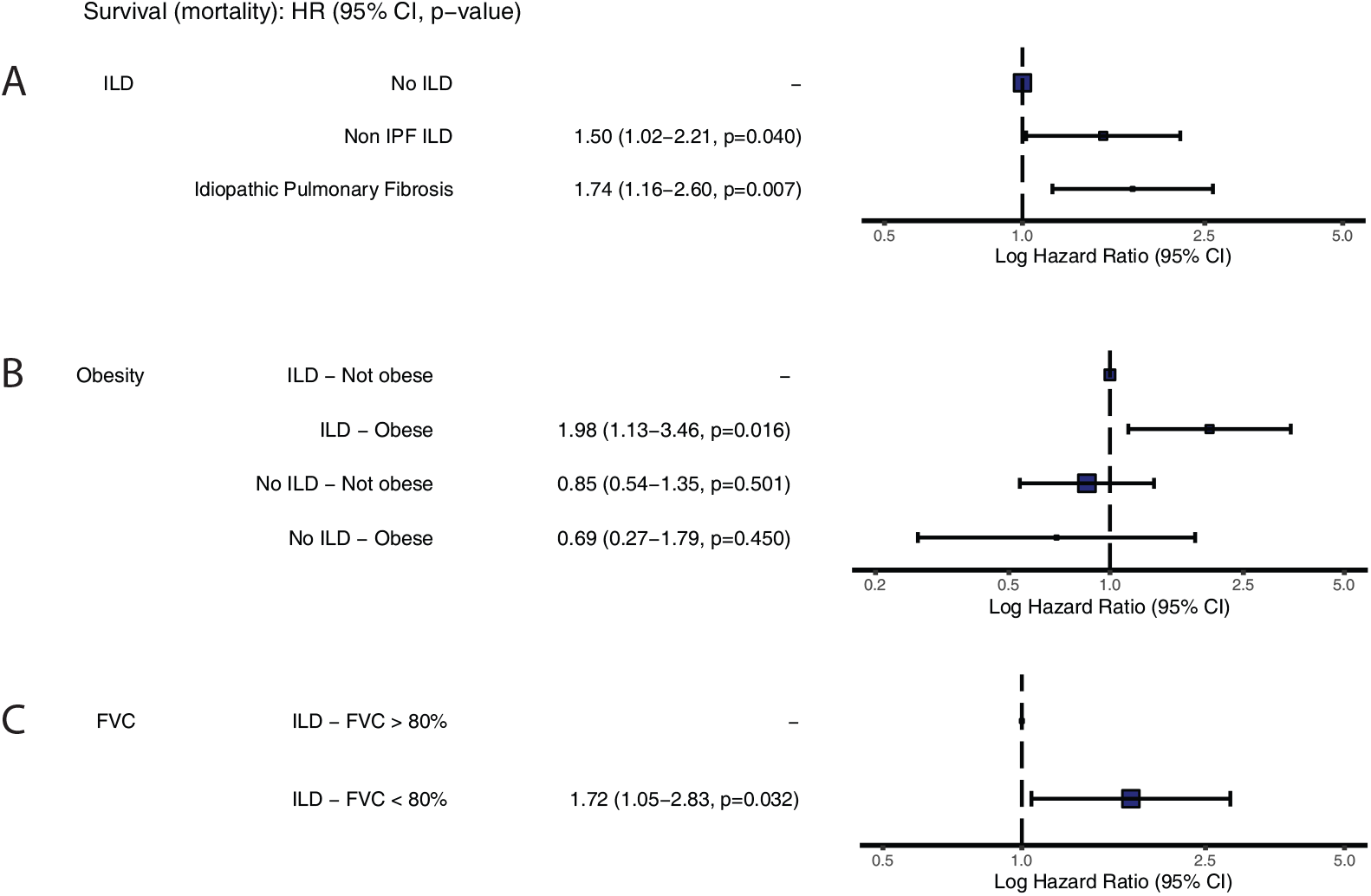
A) Effect of ILD on mortality following COVID-19 (Propensity Matched), B) effect of obesity and ILD (Propensity Matched) C) and effect of FVC on outcome in ILD patients only.

### Predictors of outcome from COVID-19

Obesity has been previously described to increase the risk of death from COVID-19. After obesity was included in the propensity score matching (129/161 ILD patients with data available, matched to 260 non-ILD patients), being obese and having ILD was significantly associated with an increased hazard of death from COVID-19 (adjusted HR 1.98, 95%CI 1.13 to 3.46, p = 0.016; figure 3b), which was greater than the effect of obesity in non-ILD patients.

To determine whether patients with more severe ILD had a greater risk of mortality with COVID-19, the last available lung function results prior to hospital admission were analysed. For all ILD patients, the mean Forced Vital Capacity (FVC) in patients surviving COVID-19 was 82.2% predicted compared with 76.8% predicted in patients who died (p=0.121). Similarly, the mean Diffusion coefficient of Carbon Monoxide (DLco) was 56.4% predicted in survivors compared with 49.6% in patients dying with COVID (p=0.072; Table 3). When the FVC was dichotomized using the 80% predicted threshold between mild and moderate ILD, in line with UK prescribing policy for anti-fibrotic medication, the risk of mortality was significantly higher in patients with moderate or severe ILD (HR 1.72, 95% CI 1.05-2.83), compared with mild disease (Figure 3c).

**Table 3.** Lung function in ILD group by death

| label | Total N | Missing N | levels | Alive | Died | p |
| --- | --- | --- | --- | --- | --- | --- |
| Forced Vital Capacity | 147 | 37 | Mean (SD) | 2.7 (1.0) | 2.6 (0.9) | 0.372* |
| Forced Vital Capacity,<br>percentage of<br>predicted | 151 | 33 | Mean (SD) | 82.2 (23.0) | 76.8 (21.0) | 0.135* |
| Forced Vital Capacity,<br>at 80% of predicted | 151 | 33 | FVC < 80% | 35 (42.7) | 42 (60.9) | 0.039 |
|  |  |  | FVC > 80% | 47 (57.3) | 27 (39.1) |  |
| Forced Vital Capacity,<br>percentage of<br>predicted | 151 | 33 | <50% | 7 (6.9) | 4 (4.8) | 0.070 |
|  |  |  | 50 to 80% | 28 (27.7) | 38 (45.8) |  |
|  |  |  | 80 to 100% | 33 (32.7) | 17 (20.5) |  |
|  |  |  | Over 100% | 14 (13.9) | 10 (12.0) |  |
|  |  |  | (Missing) | 19 (18.8) | 14 (16.9) |  |
| Diffusing capacity for<br>carbon monoxide | 81 | 103 | Mean (SD) | 5.2 (3.4) | 4.2 (1.9) | 0.093* |
| Diffusing capacity for<br>carbon monoxide,<br>percentage of<br>predicted | 122 | 62 | Mean (SD) | 56.4 (19.4) | 49.6 (21.6) | 0.072* |

SD – Standard Deviation. All tests Welch's two sample t-test, unless indicated by \* where Kruskal Wallis used.

### Ventilatory support for ILD patients with COVID-19

Most patients with ILD (84%; 135/161) did not receive enhanced respiratory support which was similar to matched patients without ILD (79%; 254/322; Table 2). Significantly more patients who survived did not receive enhanced respiratory support (93%, 76/82) compared with those who died (75%; 59/79 p=0.015). Of the 26 patients receiving enhanced respiratory support 77% (20) died, including 83% (5/6) of matched patients with ILD receiving ventilation (Table 4).

**Table 4:** Use of Ventilation in patients with ILD by outcome

|  | Total N | Missing N | levels | Alive | Died | p |
| --- | --- | --- | --- | --- | --- | --- |
| Respiratory Support | 161 | 0 | Unenhanced respiratory support* | 76 (92.7) | 59 (74.7) | 0.015 |
|  |  |  | Enhanced Respiratory Support | 5 (6.0) | 15 (23) | 0.005 |
|  |  |  | Ventilation | 1 (1.2) | 5 (3.8) | 0.190 |

All test Chi-square. \*Standard flow-rate Oxygen supplementation not considered 'enhanced' respiratory support, CPAP and High Flow O<sub>2</sub> considered enhanced respiratory support. Note a further 10 patients with ILD were ventilated but not included in the matching, of those 6 survived.

### Effect of anti-fibrotic and immunosuppressive therapy on outcome from COVID-19

In patients with ILDs 106 were taking no immunosuppressants or anti-fibrotic of whom 50% died (53/106; Table 5). Almost a third of patients with IPF (20/68) were receiving anti-fibrotic therapy at the time of admission. Seven patients were receiving pirfenidone and 13 were receiving nintedanib. Of those receiving anti-fibrotic therapy, 50% (10/20) died. Where immunosuppressants were reported, corticosteroids were most frequently prescribed (45/55) followed by mycophenolate (17/55); overall 47% (26/55) of patients taking anti-fibrotics or immunosuppressants died. Significantly more patients with ILD received oral corticosteroids in hospital than patients without ILD (27.8% versus 9.9%; p<0.001) but there was no difference in outcome between those receiving corticosteroids (48.9% mortality (22/55)) and those who did not (48.7% mortality (57/117)).

**Table 5:** Drugs received by patients with ILD

|  | levels | Alive | Died |
| --- | --- | --- | --- |
| Received Immuno-suppressants/ Anti-fibrotics | No | 53 (64.6) | 53 (67.1) |
|  | Yes | 29 (34.9) | 26 (32.9) |
| Methotrexate/ Azathioprine | No | 80 (96.4) | 77 (98.7) |
|  | Yes | 3 (3.6) | 2 (1.3) |
| Nintedanib | No | 74 (89.2) | 75 (94.9) |
|  | Yes | 9 (10.8) | 4 (5.1) |
| Pirfenidone* | No | 82 (98.8) | 73 (92.4) |
|  | Yes | 1 (1.2) | 6 (7.6) |
| Mycophenolate | No | 73 (88.0) | 72 (91.1) |
|  | Yes | 10 (12.0) | 7 (8.9) |
| Corticosteroid | No | 60 (72.3) | 57 (72.2) |
|  | Yes | 23 (27.7) | 22 (27.8) |

\*2 further patients received pirfenidone and survived but were not included in matched cohort.

## Discussion

Determining the risk of poor outcome in patients pre-existing conditions who acquire SARS-CoV-2 infection is crucial to determine what mitigation measures are required in the community. Our study shows that patients with existing interstitial lung disease were at very high risk of death when hospitalised with COVID-19, especially if they have reduced prior lung function or obesity. Most died without being offered mechanical ventilation; in those who were ventilated, mortality was very high.

Known risk factors for poor outcomes in COVID-19 include increasing age, male sex, DM, HT, IHD and COPD (9,12). Unfortunately, the risk for patients with ILD has not yet been quantified because patients with uncommon diseases often do not have their specific diagnostic information recorded in large observational studies or are often not accurately reported in administrative healthcare datasets. Despite the unknown risk of COVID-19 for patients with ILD, many patients chose to self-isolate and, therefore, there appeared to be apparent ‘protection’ from COVID-19 observed to many ILD physicians around the world. To accurately understand the risk of COVID-19 and inform decision- and risk-matrices going forward, the European ILD community undertook an audit of hospitalised patients with ILD between the 1st March and 1st May 2020 to coincide with the first peak of COVID-19 within Europe. These data represent the largest assessment of the impact of SARS-CoV-2 infection on patients with ILD to date. They demonstrate that patients with ILD are at increased risk of death following hospitalisation for COVID-19 particularly elderly males, with poor lung and obesity.

The role of viral infection promoting acute exacerbations of ILD has been investigated for a number of years without a definitive answer, which probably reflects the range of viruses studied, the number of patients needed to explore such a hypothesis, and more broadly the challenges studying acute exacerbations of ILD (13,14,15,16). These data confirm that patients with ILD are at increased risk of mortality from COVID-19 compared with a matched population without ILD and the risk is greatest for IPF consistent with a respiratory virus inducing acute exacerbations of ILD.

Factors associated with poor prognosis in acute exacerbations of ILD include pulmonary fibrosis and poor lung function prior to admission and fibrotic ILDs (17,18,19,20). The data presented here, support these prior studies demonstrating that patients with a recorded FVC <80% predicted prior to admission had an increased mortality compared with those with an FVC >80%. Similarly, patients with fibrotic ILDs such as Chronic Hypersensitivity Pneumonitis had mortality rates comparable with IPF and higher than those with Sarcoid or Connective Tissue Disease associated ILD. Interestingly, obese patients with ILD had a substantially increased risk of mortality. This may reflect use of steroids in severe disease, although we do not favour this hypothesis as there was no apparent increased risk of death associated with corticosteroid use. Obesity may be due to limited activity in ILD patients, a condition that leads to progressive reduction in exercise tolerance as it becomes more severe (21). Alternatively, and perhaps most intriguingly, it may lend support to the hypothesis that severe ILD is part of the ‘metabolic syndrome’ (22).

This analysis also assessed respiratory support offered to patients hospitalised with ILD. In line with current practice and guidelines most patients did not receive any ventilatory support (7, 23). Most patients who survived did not require enhanced respiratory support such as High Flow Oxygen, Continuous Positive Airway Pressure or ventilation, and of those receiving enhanced respiratory support the majority died. These data continue to support the use of supportive care in preference to either non-invasive or invasive mechanical ventilation except in clearly defined cases such as predominantly inflammatory ILD or bridging to transplantation.

This analysis has a number of strengths. It is the largest, international, multicentre study to assess the outcome of patients with Interstitial Lung Disease hospitalised with the respiratory infection SARS-Cov-2. Merging of the ILD Audit dataset with the ISARIC4C CCP-UK dataset enabled accurate, contemporary propensity-score matching for a number of potential confounding factors to assess the risk from COVID-19 disease. This enabled an assessment of the risk to patients with ILD infected with SARS-CoV-2, facilitating evidence-based mitigation strategies, such as self-isolation for this vulnerable group of patients. This study also has some potential weaknesses. Due to the retrospective nature of the data collection recall bias might have led to over-selection of severe cases of COVID-19 in patients with ILD, however, this is mitigated by the large number of centres participating in the audit. Similarly, because only hospitalised patients could be included, it is possible that a large number of patients with ILD and COVID-19 were omitted and, therefore, the risk of COVID-19 could be over-stated. However, given the demographic associated with ILD we think this is unlikely. Also, the propensity-score matching included some younger patients who would be expected to have less severe disease, although there was insufficient detail in the audit data collection to match for COVID-19 severity based on admission severity scores. The effect of obesity was assessed in the matched population suggesting an increased risk in patients without ILD, however in contrast with prior reports (9,24) no effect of obesity was observed in the control population. This probably reflects the relatively small numbers of patients used in the matching and the large amount of missing data relating to weight. Therefore, the data relating to obesity must be interpreted accordingly, but we believe as a modifiable risk factor it is important to highlight the risk of obesity in patients with ILD who might develop COVID-19. Finally, it was not possible to evaluate specific treatment effects, such as the use of anti-fibrotics and immunosuppressants. Only a proportion of patients received anti-fibrotics during their admission. Furthermore, due to the unknown effect of anti-fibrotics, and concerns regarding immunosuppression on the clinical course of COVID-19 there is likely to be considerable confounding due to indication. There are both potential benefits (25,26) and harms associated with background therapy for ILD (27). However, it is reassuring from a safety perspective that there was no obvious signal suggesting harms associated with therapy for ILD in patients with COVID-19, although, further study to evaluate the consequence of anti-fibrotic and immunomodulatory therapy in ILD are needed.

In summary, these data demonstrate that patients with ILD, particularly those with fibrotic ILD are at higher risk of mortality from COVID-19 than patients without ILD. Furthermore, the risk is heightened in elderly males, those with obesity or poor lung function and we would recommend dietary advice in overweight patients. Ventilatory support did not objectively improve outcomes for COVID-19 and we therefore suggest that ventilatory support only be offered in select subgroups of patients in line with current recommendations. Similarly, anti-fibrotic, or immunosuppressive medication did not objectively alter outcome and therefore we do not propose these medications should be withheld during future COVID-19 surges. We also propose that patients with ILD, particularly severe fibrotic ILD, continue to be regarded as high risk of mortality from COVID-19 and follow national self-isolation guidelines for vulnerable individuals and be prioritised for SARS-CoV-2 vaccination at such time as it becomes available. Finally, we believe these data demonstrate the importance of international collaboration to collect data to understand the consequences of emerging threats to patients with ILD.

## Data Availability

All de-identified, anonymised, and unlinked data from the ILD Audit can be shared on request. ISARIC 4C data can be accessed in line with data sharing policy

https://isaric4c.net

## ISARIC 4C Investigators

Consortium Lead Investigator J Kenneth Baillie, Chief Investigator Malcolm G Semple Co-Lead Investigator Peter JM Openshaw. ISARIC Clinical Coordinator Gail Carson. Co-Investigators: Beatrice Alex, Benjamin Bach, Wendy S Barclay, Debby Bogaert, Meera Chand, Graham S Cooke, Annemarie B Docherty, Jake Dunning, Ana da Silva Filipe, Tom Fletcher, Christopher A Green, Ewen M Harrison, Julian A Hiscox, Antonia Ying Wai Ho, Peter W Horby, Samreen Ijaz, Saye Khoo, Paul Klenerman, Andrew Law, Wei Shen Lim, Alexander, J Mentzer, Laura Merson, Alison M Meynert, Mahdad Noursadeghi, Shona C Moore, Massimo Palmarini, William A Paxton, Georgios Pollakis, Nicholas Price, Andrew Rambaut, David L Robertson, Clark D Russell, Vanessa Sancho-Shimizu, Janet T Scott, Louise Sigfrid, Tom Solomon, Shiranee Sriskandan, David Stuart, Charlotte Summers, Richard S Tedder, Emma C Thomson, Ryan S Thwaites, Lance CW Turtle, Maria Zambon. Project Managers Hayley Hardwick, Chloe Donohue, Jane Ewins, Wilna Oosthuyzen, Fiona Griffiths. Data Analysts: Lisa Norman, Riinu Pius, Tom M Drake, Cameron J Fairfield, Stephen Knight, Kenneth A Mclean, Derek Murphy, Catherine A Shaw. Data and Information System Manager: Jo Dalton, Michelle Girvan, Egle Saviciute, Stephanie Roberts Janet Harrison, Laura Marsh, Marie Connor. Data integration and presentation: Gary Leeming, Andrew Law, Ross Hendry. Material Management: William Greenhalf, Victoria Shaw, Sarah McDonald. Local Principal Investigators: Kayode Adeniji, Daniel Agranoff, Ken Agwuh, Dhiraj Ail, Ana Alegria, Brian Angus, Abdul Ashish, Dougal Atkinson, Shahedal Bari, Gavin Barlow, Stella Barnass, Nicholas Barrett, Christopher Bassford, David Baxter, Michael Beadsworth, Jolanta Bernatoniene, John Berridge, Nicola Best, Pieter Bothma, David Brealey, Robin Brittain-Long, Naomi Bulteel, Tom Burden, Andrew Burtenshaw, Vikki Caruth, David Chadwick, Duncan Chambler, Nigel Chee, Jenny Child, Srikanth Chukkambotla, Tom Clark, Paul Collini, Catherine Cosgrove, Jason Cupitt, Maria-Teresa Cutino-Moguel, Paul Dark, Chris Dawson, Samir Dervisevic, Phil Donnison, Sam Douthwaite, Ingrid DuRand, Ahilanadan Dushianthan, Tristan Dyer, Cariad Evans, Chi Eziefula, Chrisopher Fegan, Adam Finn, Duncan Fullerton, Sanjeev Garg, Sanjeev Garg, Atul Garg, Jo Godden, Arthur Goldsmith, Clive Graham, Elaine Hardy, Stuart Hartshorn, Daniel Harvey, Peter Havalda, Daniel B Hawcutt, Maria Hobrok, Luke Hodgson, Anita Holme, Anil Hormis, Michael Jacobs, Susan Jain, Paul Jennings, Agilan Kaliappan, Vidya Kasipandian, Stephen Kegg, Michael Kelsey, Jason Kendall, Caroline Kerrison, Ian Kerslake, Oliver Koch, Gouri Koduri, George Koshy, Shondipon Laha, Susan Larkin, Tamas Leiner, Patrick Lillie, James Limb, Vanessa Linnett, Jeff Little, Michael MacMahon, Emily MacNaughton, Ravish Mankregod, Huw Masson, Elijah Matovu, Katherine McCullough, Ruth McEwen, Manjula Meda, Gary Mills, Jane Minton, Mariyam Mirfenderesky, Kavya Mohandas, Quen Mok, James Moon, Elinoor Moore, Patrick Morgan, Craig Morris, Katherine Mortimore, Samuel Moses, Mbiye Mpenge, Rohinton Mulla, Michael Murphy, Megan Nagel, Thapas Nagarajan, Mark Nelson, Igor Otahal, Mark Pais, Selva Panchatsharam, Hassan Paraiso, Brij Patel, Justin Pepperell, Mark Peters, Mandeep Phull, Stefania Pintus, Jagtur Singh Pooni, Frank Post, David Price, Rachel Prout, Nikolas Rae, Henrik Reschreiter, Tim Reynolds, Neil Richardson, Mark Roberts, Devender Roberts, Alistair Rose, Guy Rousseau, Brendan Ryan, Taranprit Saluja, Aarti Shah, Prad Shanmuga, Anil Sharma, Anna Shawcross, Jeremy Sizer, Richard Smith, Catherine Snelson, Nick Spittle, Nikki Staines, Tom Stambach, Richard Stewart, Pradeep Subudhi, Tamas Szakmany, Kate Tatham, Jo Thomas, Chris Thompson, Robert Thompson, Ascanio Tridente, Darell Tupper - Carey, Mary Twagira, Andrew Ustianowski, Nick Vallotton, Lisa Vincent-Smith, Shico Visuvanathan, Alan Vuylsteke, Sam Waddy, Rachel Wake, Andrew Walden, Ingeborg Welters, Tony Whitehouse, Paul Whittaker, Ashley Whittington, Meme Wijesinghe, Martin Williams, Lawrence Wilson, Sarah Wilson, Stephen Winchester, Martin Wiselka, Adam Wolverson, Daniel G Wooton, Andrew Workman, Bryan Yates, Peter Young.

## Acknowledgements

This work uses data provided by patients and collected by the NHS as part of their care and support #DataSavesLives. We are extremely grateful to the 2,648 frontline NHS clinical and research staff and volunteer medical students, who collected this data in challenging circumstances; and the generosity of the participants and their families for their individual contributions in these difficult times. We also acknowledge the support of Jeremy J Farrar, Nahoko Shindo, Devika Dixit, Nipunie Rajapakse, Piero Olliaro, Lyndsey Castle, Martha Buckley, Debbie Malden, Katherine Newell, Kwame O’Neill, Emmanuelle Denis, Claire Petersen, Scott Mullaney, Sue MacFarlane, Chris Jones, Nicole Maziere, Katie Bullock, Emily Cass, William Reynolds, Milton Ashworth, Ben Catterall, Louise Cooper, Terry Foster, Paul Matthew Ridley, Anthony Evans, Catherine Hartley, Chris Dunn, D. Sales, Diane Latawiec, Erwan Trochu, Eve Wilcock, Innocent Gerald Asiimwe, Isabel Garcia-Dorival, J. Eunice Zhang, Jack Pilgrim, Jane A Armstrong, Jordan J. Clark, Jordan Thomas, Katharine King, Katie Alexandra Ahmed, Krishanthi S Subramaniam, Lauren Lett, Laurence McEvoy, Libby van Tonder, Lucia Alicia Livoti, Nahida S Miah, Rebecca K. Shears, Rebecca Louise Jensen, Rebekah Penrice-Randal, Robyn Kiy, Samantha Leanne Barlow, Shadia Khandaker, Soeren Metelmann, Tessa Prince, Trevor R Jones, Benjamin Brennan, Agnieska Szemiel, Siddharth Bakshi, Daniella Lefteri, Maria Mancini, Julien Martinez, Angela Elliott, Joyce Mitchell, John McLauchlan, Aislynn Taggart, Oslem Dincarslan, Annette Lake, Claire Petersen, Scott Mullaney and Graham Cooke. We would like to thank Lise-Ann Grundy, Mark Major and Amanda Bell who collected data.

Outcome of COVID-19 in patients hospitalised with Interstitial Lung Disease: An international multicentre study. Supplementary Material.

---

**Supplementary Table 1.**

| label | 'Other' ILD diagnoses | n |
| --- | --- | --- |
| Other ILD Details | Age-related ILD/presbyotic lung | <3 |
|  | ANCA vasculitis | <3 |
|  | asbestosis | 5 |
|  | Autoimmune Pneumonitis | 5 |
|  | Chronic eosinophilic pneumonia | <3 |
|  | Combined Emphysema and Pulmonary Fibrosis | 7 |
|  | Desquamative Intestinal Pneumonia | <3 |
|  | Non-Specific Interstitial Pneumonia | 10 |
|  | Organising Pneumonia | <3 |
|  | PPFE | <3 |
|  | smoking related ILD | <3 |
|  | Unclassifiable ILD | 11 |

**Figure S1.**
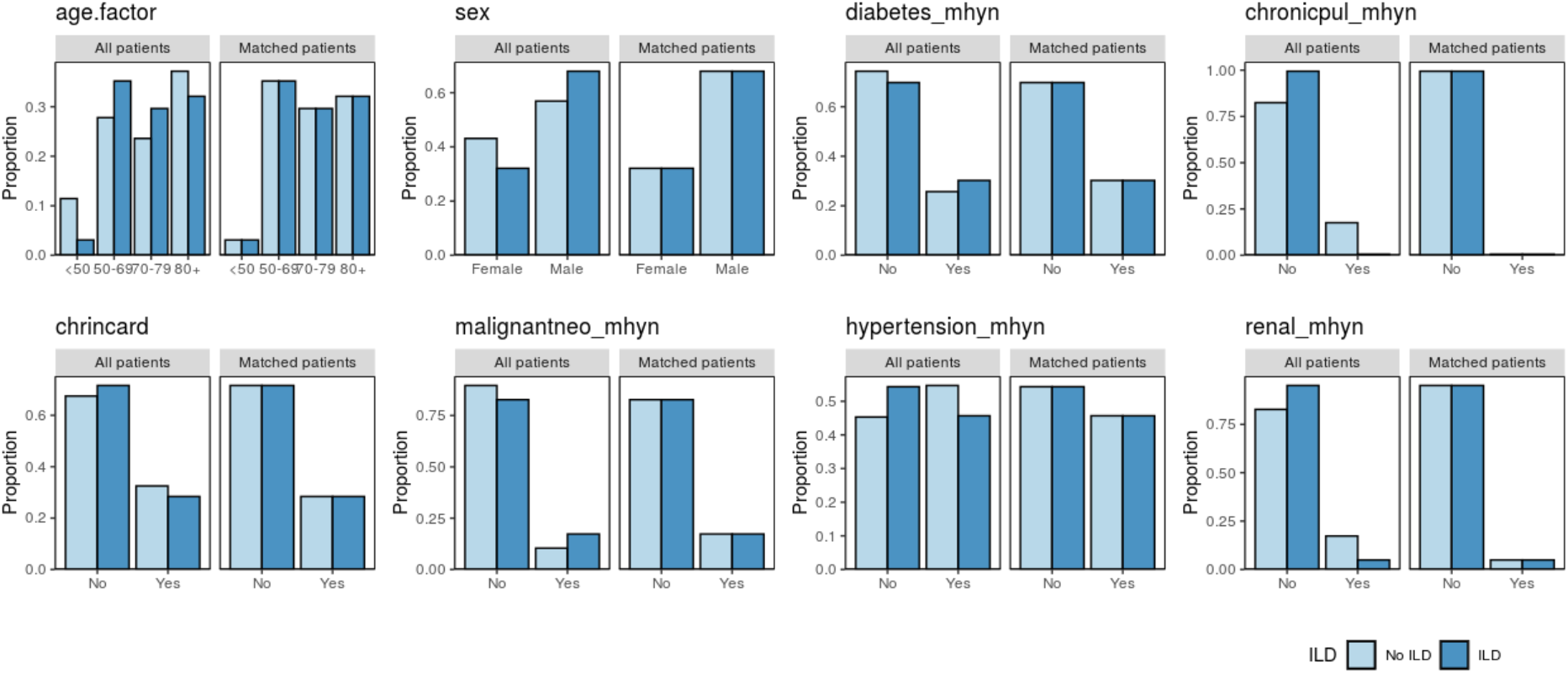
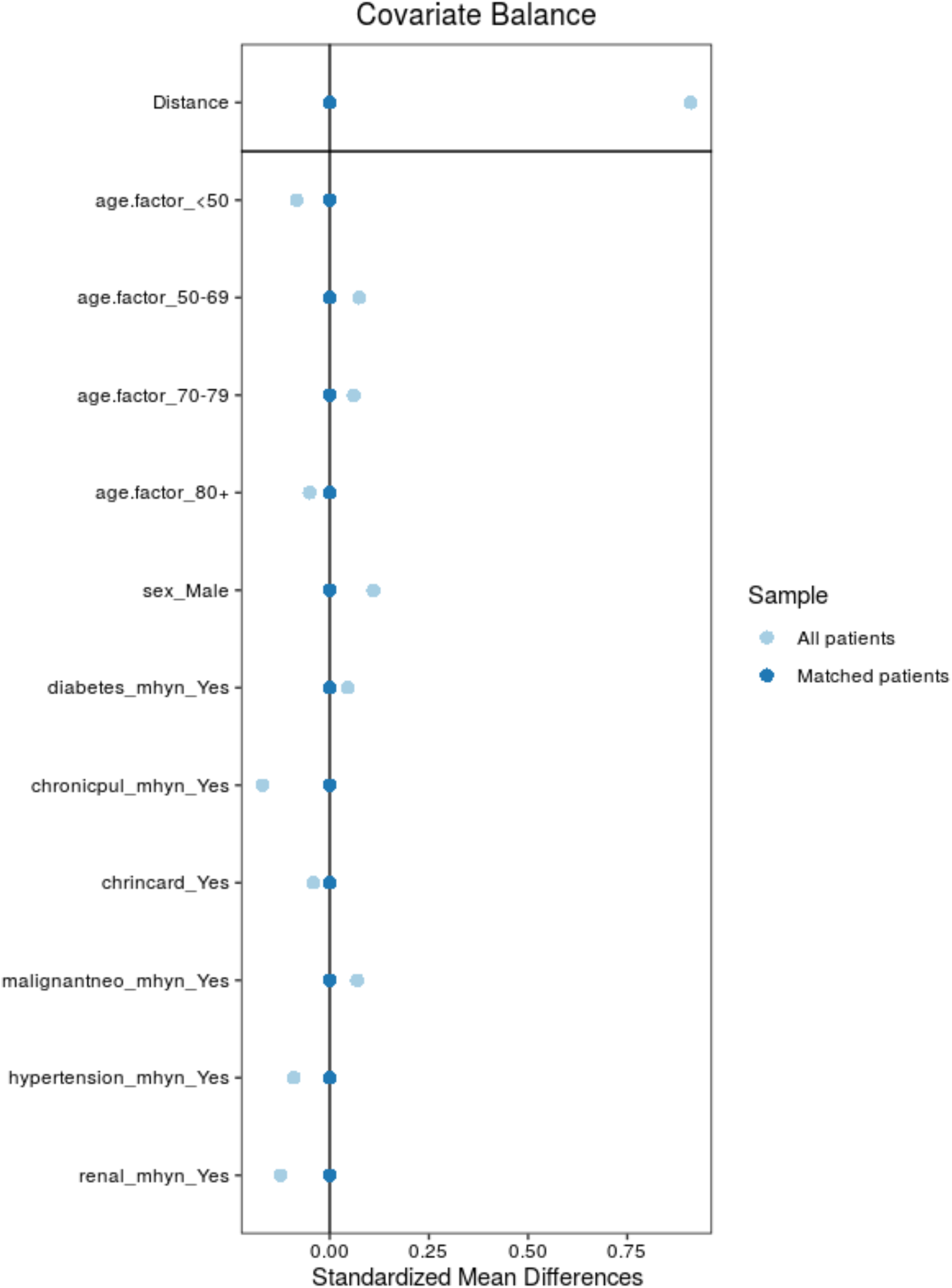
Balance Plots. Distribution of differences across ILD groups by matching variable. Propensity score matching normalised standardised mean differences across the treatment groups, with observed differences before matching not being present after.

**Figure S2.**
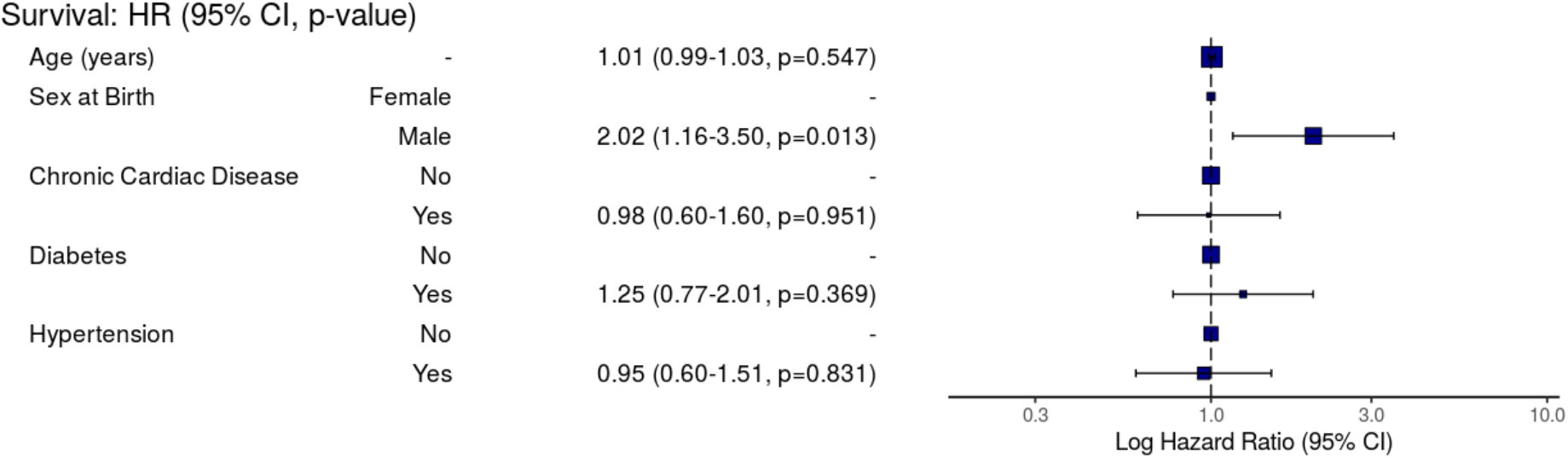
Impact of sex, and co-morbidity on outcomes of COVID-19 in ILD

